# Hexosylceramides and glycerophosphatidylcholine GPC(36:1) increase in Multi-Organ Dysfunction Syndrome patients with Pediatric Intensive Care Unit Admission over 8-day hospitalization

**DOI:** 10.1101/2021.02.11.21251596

**Authors:** Mara L. Leimanis-Laurens, Emily Wolfrum, Karen Ferguson, Jocelyn R. Grunwell, Dominic Sanfilippo, Jeremy W Prokop, Todd A. Lydic, Surender Rajasekaran

**Affiliations:** Pediatric Critical Care Unit, Helen DeVos Children’s Hospital, Grand Rapids, MI, 49503, United States.; Department of Pediatric and Human Development, College of Human Medicine, Michigan State University, Life Sciences Bldg. 1355 Bogue Street, East Lansing MI 48824.; Bioinformatics & Biostatistics Core, Van Andel Institute, Grand Rapids, MI, United States.; Emory University & Children’s Healthcare of Atlanta, Atlanta, GA, 30322 United States.; Department of Pharmacology and Toxicology, Michigan State University, East Lansing, MI 48824 USA.; Collaborative Mass Spectrometry Core Department of Physiology, East Lansing, MI, United States.

**Keywords:** lipidomics, pediatrics, critical illness, multi-organ dysfunction syndrome, glycerolipids, glycerophosphatidylcholine, sphingolipids, sphingomyelin

## Abstract

Glycero- and sphingo-lipids are important in plasma membrane structure, caloric storage and signaling. An un-targeted lipidomics approach for a cohort of critically ill pediatric intensive care unit (PICU) patients, undergoing multi-organ dysfunction syndrome (MODS) was compared to sedation controls. After IRB approval, patients meeting criteria for MODS were screened, consented (n=24), and blood samples were collected from the PICU at HDVCH, Michigan; eight patients needed veno-arterial extracorporeal membrane oxygenation (VA ECMO). Sedation controls were presenting for routine sedation (n=4). Plasma lipid profiles were determined by nano-electrospray (nESI) direct infusion high resolution/accurate mass spectrometry (MS) and tandem mass spectrometry (MS/MS). Biostatistics analysis was performed using R v 3.6.0. 61 patient samples over 3 time points revealed a ceramide metabolite, hexosylceramide (Hex-Cer) was high across all time points (mean 1.63% - 3.19%; vs. controls 0.22%). Fourteen species statistically differentiated from sedation controls (P-value ≤0.05); sphingomyelin (SM) [SM(d18:1/23:0), SM(d18:1/22:0), SM(d18:1/23:1), SM(d18:1/21:0), SM(d18:1/24:0)]; and glycerophosphotidylcholine (GPC) [GPC(36:01), GPC(18:00), GPC(O:34:02), GPC(18:02), GPC(38:05), GPC(O:34:03), GPC(16:00), GPC(40:05), GPC(O:36:03)]. Hex-Cer has been shown to be involved in viral infection and may be at play during acute illness. GPC(36:01) was elevated in all MODS patients at all time points and is associated with inflammation and brain injury.

## 1. Introduction

The human plasma lipidome has been well described in the last decade [1] with the launch of the Lipid Maps Consortium (www.lipidmaps.org). From this work it has been found that over 600 unique molecular species exist, with a further estimate in the hundreds of thousands [2], half of which consist of glycerolipids, glycerophospholipids and sphingolipids. All of these lipid classes are structurally constituted of a fatty acid with long hydrocarbon chains, and a glycerol group, with sphingolipids as the only exception lacking a glycerol group (See Figure 1; from Quehenberger and Dennis [1]).

### 1.1 Multi-organ dysfunction syndrome (MODS) and role of lipids in critical illness

Multi-organ dysfunction syndrome (MODS) has been previously described as a major source of pediatric intensive care unit (PICU) admissions [3], and fatalities [4]. A portion of the patients with MODS are due to infections, as most of our current understanding of the pathophysiology has been taken from work done in pediatric sepsis [5–8]. There exist a smaller cohort of patients that requires aggressive life support measures such as extracorporeal membrane oxygenation (ECMO), with 45% reported mortality [9].

More recently, in the nascent field of lipidomics, effects of infection from viruses has been described in plasma, namely for COVID-19 [10], and Ebola [11]. As the specific assault on the patient appears to harbor different metabolic consequences for its victims. Perhaps the most well characterized and described of all the lipids are plasma glycer-olipids, namely triacyglycerides (TAGs), which are famously involved in many metabolic syndromes [12,13], along with diacylglycerols (DAGs) [14]. TAG’s have been found to be elevated in children with systemic inflammatory response syndrome and sepsis [12] and in adults with acute respiratory distress syndrome [13].

Sphingolipids have been characterized in infection, autoimmunity and neurodegenerative disorders. Sphingolipids and the sphingolipidome have been recently characterized in the involvement in hemophagocytic lymphohistiocytosis [15], linked to Alzheimer’s [16], and inflammation [17]. Given the existing literature we anticipate seeing differences in these lipid classes in our patients.

### 1.2 Lipidomics as Biomarkers in critical illness

Little is known on the dyslipemia during the height of critical illness. From a previous report [18] we found that phospholipids were highly down-regulated in the acute phase of illness, as patients were being admitted to the PICU, and partially recovered with the introduction of feeds, by the 8-day mark. Based on these initial findings, we have expanded our analysis to further evaluate potential effects of critical illness on a broader range of lipids, including glycerolipids, glycerophosphatidylcholine, sphingolipids and sphingomyelins over an 8-day PICU course.

## 2. Results

### 2.1 Study Description and Demographics

Basic demographics and dietary intake were previously reported [18,19]. In brief, we had a total of 61 patient samples over 3 time points for 28 patients. All patients were being mechanically ventilated at the time of the baseline sample collection and receiving inotropes. Additional organs affected were kidneys (n=15; 54 %), including 7 MODS and all 8 ECMO patients (statistical test, p-value=0.01); liver (n=8; 28.6 %; 4 MODS; 4 ECMO), and brain (n=6; 21.4 %; 4 MODS; 2 ECMO). Severity of illness scores Pediatric Logistic Organ Dysfunction-2 score (PELOD) were not significantly different amongst groups across time points (Global F-stat, p=0.66).

### 2.2 Percent total lipids over time and treatment course

#### 2.2.1 Sphingolipids and Glycerolipids

Lipid percentages for glycerolipids (including TAG’s) and sphingolipids are listed in Table 1. Phospholipids constitute the vast majority of plasma lipids and were described in a previous report [18]. The second major lipid group in terms of total percentage include the sphingolipids with mean value of 8% for MODS at baseline, which is much lower than 13% for sedation controls (t-test, p=0.0056). By 8 days the mean value is 15% for the MODS group, indicating a dramatic doubling over the 8-day course. ECMO patients demonstrated an intermediate profile. Glycerolipid values in all patients were higher than sedation controls by day 8, and are typically the major lipid in plasma. Sphingolipids are low at baseline and increase over time, the glycerolipids are higher already at baseline and go at 8 days.

**Table 1.** Percent total of major lipid classes of MODS/ECMO Study 2016-2018 (*N*=28)

| Lipid Class | Sphingolipids |  |  |  | Glycerolipids |  |  |  |
| --- | --- | --- | --- | --- | --- | --- | --- | --- |
|  | Mean | SD | Median | P-value | Mean | SD | Median | P-value |
| **Sedation Controls (n=4)** | 13.03 | 1.24 | 13.21 |  | 3.43 | 3.34 | 2.57 |  |
| **MODS BL (n=16)** | 7.86 | 3.18 | 7.40 | **P = 0.0056** | 12.25 | 13.35 | 5.39 | P = 0.0296* |
| **ECMO BL (n=8)** | 12.32 | 2.59 | 11.91 | P = 0.6217 | 10.10 | 8.63 | 8.37 | P = 0.1746 |
| **MODS 72h (n=15)** | 9.06 | 3.82 | 9.88 | P = 0.0601 | 9.58 | 7.16 | 6.75 | P = 0.1182 |
| **ECMO 72h (n=7)** | 11.50 | 3.22 | 10.80 | P = 0.3952 | 8.51 | 9.20 | 4.19 | P = 0.3234 |
| **MODS 8d (n=8)** | 14.85 | 3.71 | 15.20 | P = 0.3713 | 10.01 | 6.10 | 9.59 | P = 0.0759 |
| **ECMO 8d (n=6)** | 11.36 | 2.33 | 11.16 | P = 0.2305 | 11.67 | 5.48 | 11.80 | P = 0.0285 |

Notes: All tests are using sedation as comparative group; T-test performed (assuming equal variances); \*F-test for equal variances was $P < 0.05$ Welch-test (assuming unequal variances) was used. Using Bonferroni correction for multiple comparisons, adjusted for $P < 0.0083$ ; BL: baseline; 72h: 72 hours; 8d: 8 days.

#### 2.2.2 Sub-groups of sphingolipids

From these initial observations of total plasma sphingolipids we were able to quantify structurally varied groups, namely dihydrosphingomyelin (dhSM), sphingomyelin (SM), ceramides (Cer), 2-hydroxy Ceramides (2-hydroxy Cer), Hexosylceramides (Hex-Cer), lactosylceramides (Lac-Cer) (Table 2). The overwhelming majority of the sphingolipids were SM’s with median ranges in sick patients from 95% compared to 99.8% in our sedation control patients. All of the MODS patients had significantly less SM’s as compared to sedation controls when analyzed using a t-test over the 3 time points with p-values ranging from <0.0023 - <0.0001. This is likely what is contributing to the low levels of sphingosine. Both dhSM and 2-hydroxy Cer had below detectable limits in the sedation controls which made the comparatives impossible. DhSM which has been shown to contribute to membrane rigidity inhibiting viral-cell membrane fusion [20], was detected in ECMO patients only at all three time points with mean ranges of 0.75-1.39%, the maximum level was achieved at 8 days. 2-hydroxy Cer which is a precursor to ceramide and been associated with pro-apoptotic activity [21] was detected at 72 hours and 8 days with mean ranges from 0.17% to 0.48% detected per group. One metabolite of ceramide Hex-Cer was higher in the MODS patients across all time points with mean percent differences ranging from 1.63% - 3.07% vs. 0.22% (t-test, p-values: 0.003-<0.0001), values continued to increase at 8 days, at which time ECMO patients also demonstrate a statistical difference (t-test, p-value: <0.001). Lac-Cer has been recently shown to have a protective effect for type II diabetes [22], and seems to follow a similar patterning to Hex-Cer with elevated mean percent values at 72 hours for MODS patients 0.37% (t-test, p=0.0011), and ECMO patients at 8 days (t-test, p=0.0045).

**Table 2.** Percent total sphingolipids of MODS/ECMO Study 2016-2018 (*N*=28) 102

| Lipid Class | dhSM |  |  |  | SM |  |  |  | Cer |  |  |  | 2-hydroxy Cer |  |  |  | Hex-Cer |  |  |  | Lac-Cer |  |  |  |
| --- | --- | --- | --- | --- | --- | --- | --- | --- | --- | --- | --- | --- | --- | --- | --- | --- | --- | --- | --- | --- | --- | --- | --- | --- |
|  | Mean | SD | Mdn | P-value | Mean | SD | Mdn | P-value | Mean | SD | Mdn | P-value | Mean | SD | Mdn | P-value | Mean | SD | Mdn | P-value | Mean | SD | Mdn | P-value |
| **Sedation** |  |  |  |  |  |  |  |  |  |  |  |  |  |  |  |  |  |  |  |  |  |  |  |  |
| (n=4) | 0.00 | 0.00 | 0.00 |  | 99.71 | 0.25 | 99.79 |  | 2.54 | 1.52 | 2.32 |  | 0.00 | 0.00 | 0.00 |  | 0.22 | 0.25 | 0.15 |  | 0.04 | 0.03 | 0.05 |  |
| **MODS BL** |  |  |  |  |  |  |  | **P =** |  |  |  | **P =** |  |  |  |  |  |  |  | **P =** |  |  |  | **P =** |
| (n=16) | 0.00 | 0.00 | 0.00 | N/A | 95.22 | 3.98 | 96.33 | **0.0004\*** | 3.04 | 3.51 | 1.79 | 0.79 | 0.00 | 0.00 | 0.00 | N/A | 1.63 | 1.57 | 1.19 | **0.0033\*** | 0.11 | 0.10 | 0.09 | **0.0210** |
| **ECMO BL** |  |  |  |  |  |  |  | **P =** |  |  |  | **P =** |  |  |  |  |  |  |  | **P =** |  |  |  | **P =** |
| (n=8) | 0.75 | 0.62 | 1.15 | N/A | 96.66 | 2.64 | 97.39 | **0.0143\*** | 1.98 | 1.38 | 1.88 | 0.53 | 0.17 | 0.20 | 0.09 | N/A | 2.41 | 2.35 | 1.27 | **0.0347\*** | 0.15 | 0.12 | 0.09 | **0.0367\*** |
| **MODS 72h** |  |  |  |  |  |  |  | **P <** |  |  |  | **P =** |  |  |  |  |  |  |  | **P <** |  |  |  | **P =** |
| (n=15) | 0.00 | 0.00 | 0.00 | N/A | 95.17 | 1.79 | 95.50 | **0.0001\*** | 1.49 | 1.04 | 1.21 | 0.12 | 0.25 | 0.43 | 0.06 | N/A | 2.73 | 1.59 | 2.46 | **0.0001\*** | 0.37 | 0.31 | 0.31 | **0.0011\*** |
| **ECMO 72h** |  |  |  |  |  |  |  | **P =** |  |  |  | **P =** |  |  |  |  |  |  |  | **P =** |  |  |  | **P =** |
| (n=7) | 1.05 | 1.05 | 1.15 | N/A | 95.67 | 2.12 | 96.77 | **0.0025\*** | 1.02 | 0.51 | 0.85 | 0.15 | 0.41 | 0.32 | 0.56 | N/A | 3.19 | 2.65 | 1.97 | **0.0260\*** | 0.06 | 0.05 | 0.06 | 0.3609 |
| **MODS 8d** |  |  |  |  |  |  |  | **P <** |  |  |  | **P =** |  |  |  |  |  |  |  | **P =** |  |  |  | **P =** |
| (n=8) | 0.00 | 0.00 | 0.00 | N/A | 96.03 | 1.22 | 95.91 | **0.0001\*** | 0.37 | 0.25 | 0.35 | 0.07 | 0.32 | 0.27 | 0.37 | N/A | 3.07 | 1.13 | 3.18 | **0.0001\*** | 0.21 | 0.10 | 0.21 | **0.0090** |
| **ECMO 8d** |  |  |  |  |  |  |  | **P =** |  |  |  | **P =** |  |  |  |  |  |  |  | **P =** |  |  |  | **P =** |
| (n=6) | 1.39 | 1.09 | 1.49 | N/A | 95.12 | 1.95 | 94.98 | **0.0023\*** | 0.66 | 0.52 | 0.51 | 0.10 | 0.48 | 0.77 | 0.12 | N/A | 3.05 | 1.07 | 3.04 | **0.0008\*** | 0.41 | 0.18 | 0.42 | **0.0045** |

Notes: All tests are using sedation as comparative group; \*Welch-test (assuming unequal variances); BL: baseline; 72h: 72 hours; 8d: 8 days; 2-hydroxy Cer: 2-hydroxy ceramide; Cer: ceramide; dhSM: dihydrosphingomyelin; SD: standard deviation; Hex-Cer: hexosylceramides; Lac-Cer: lactosylceramides; Mdn: Median; SM: Sphingomyelin

#### 2.2.3 Lipids species for Sphingolipids and Glycerolipids at baseline and over time

We were interested to review lipids at the species level. This allowed us to determine the length of the backbone carbon-chain as well as the number of double bonds (level of saturation). Baseline samples were analyzed only, given that patients were nil per os (NPO), and nutrition intervention was minimized. A total of five SM species were found to be statistically significantly different as compared to sedation controls when analyzed using a generalized linear model, SM(d18:1/23:0; p=0.007), SM(d18:1/22:0; p=0.012), SM(d18:1/23:1; p=0.023), SM(d18:1/21:0; p=0.03), SM(d18:1/24:0; p=0.05) (Figure 2A, B).

**Figure 2:**
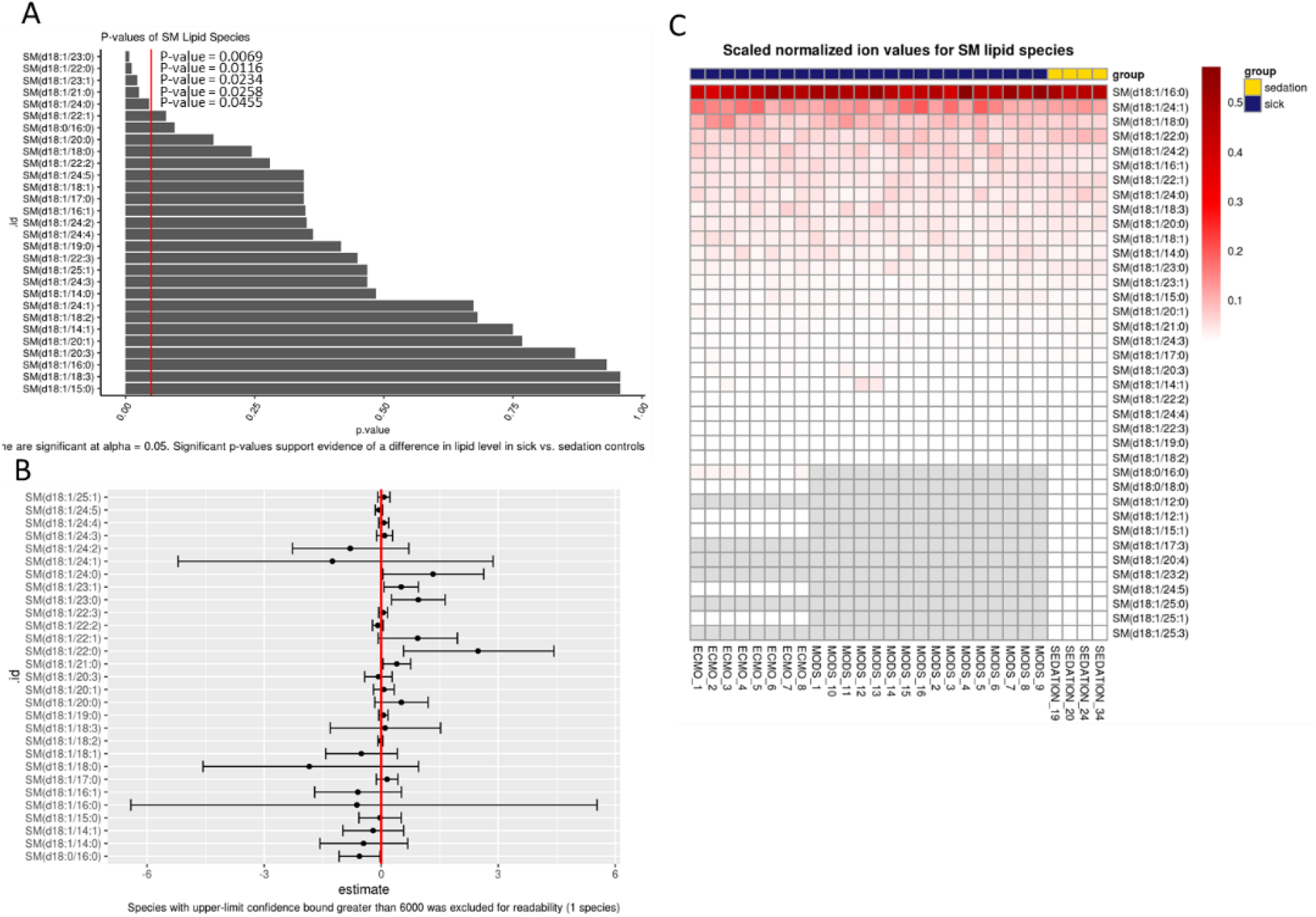
Percent total sphingomyelin lipid species at baseline.

All except one SM(d18:1/23:1) was with saturated carbon-chains, and no double bonds. The heatmap shows generally less variability in sick patients (Figure 2C).

A total of nine glycerophosphotidylcholine (GPC) species were found to be significantly different in critically ill children compared to sedation controls when analyzed with a generalized linear model GPC(36:01; p=0.002), GPC(18:00; p=0.002), GPC(O:34:02; p=0.005), GPC(18:02; p=0.006), GPC(38:05; p=0.007), GPC(O:34:03; p=0.016), GPC(16:00; p=0.020), GPC(40:05; p=0.033), GPC(O:36:03; p=0.05) (Figure 3A, B). With the exception of GPC(36:01), all of the other eight GPC species were lower than sedation controls. Choline is a marker of cellular integrity, membrane damage and turnover. The heatmap reveals a complete absence of certain species such as GPC(34:01) which implies a vast diversity of lipid species (Figure 3C).

**Figure 3.**
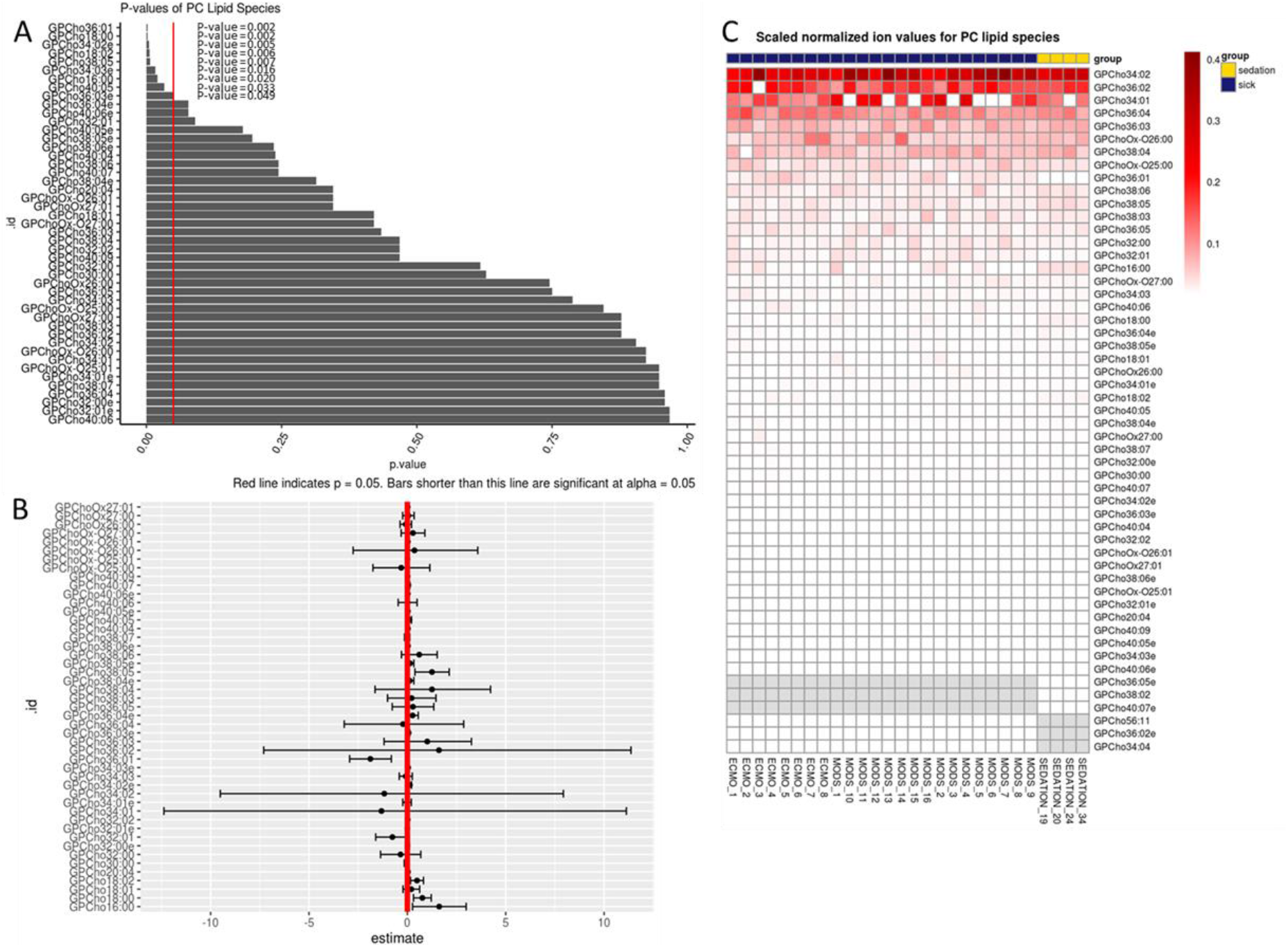
Percent total glycerophosphotidylcholine lipid species at baseline.

Additional generalized linear model analysis was run on Cholesterol (Chol) and cholesteryl esters, and TAG’s which did not reveal any statistically significant changes between the sick and sedation controls at the lipid species level, and were not further reviewed (Supplemental Figures 1,2). Diglycerides did reveal one single species (DG36:01) in the sedation control group (Supplemental Figure 3), a patterning which was lower for both MODS and ECMO patients.

Nutrition intervention on remodeling of the lipidome was determined by 72 hours for all patients. Visualization of changes in key lipid species from Figures 2 and 3 over time, are summarized in Figure 4a (as boxplots) and Figure 4b (as geometric mean). Generally, all lipid species were increasing over time. This may indicate a shift from the acute phase of illness into the recovery, as in the case of SM(d18:21:0), however the majority still remain lower as compared to sedation controls. Consequently, one GPC species GPC(36:01) which was found to be higher than sedation controls in Figure 3, is also increasing upwards over time.

**Figure 4a,b:**
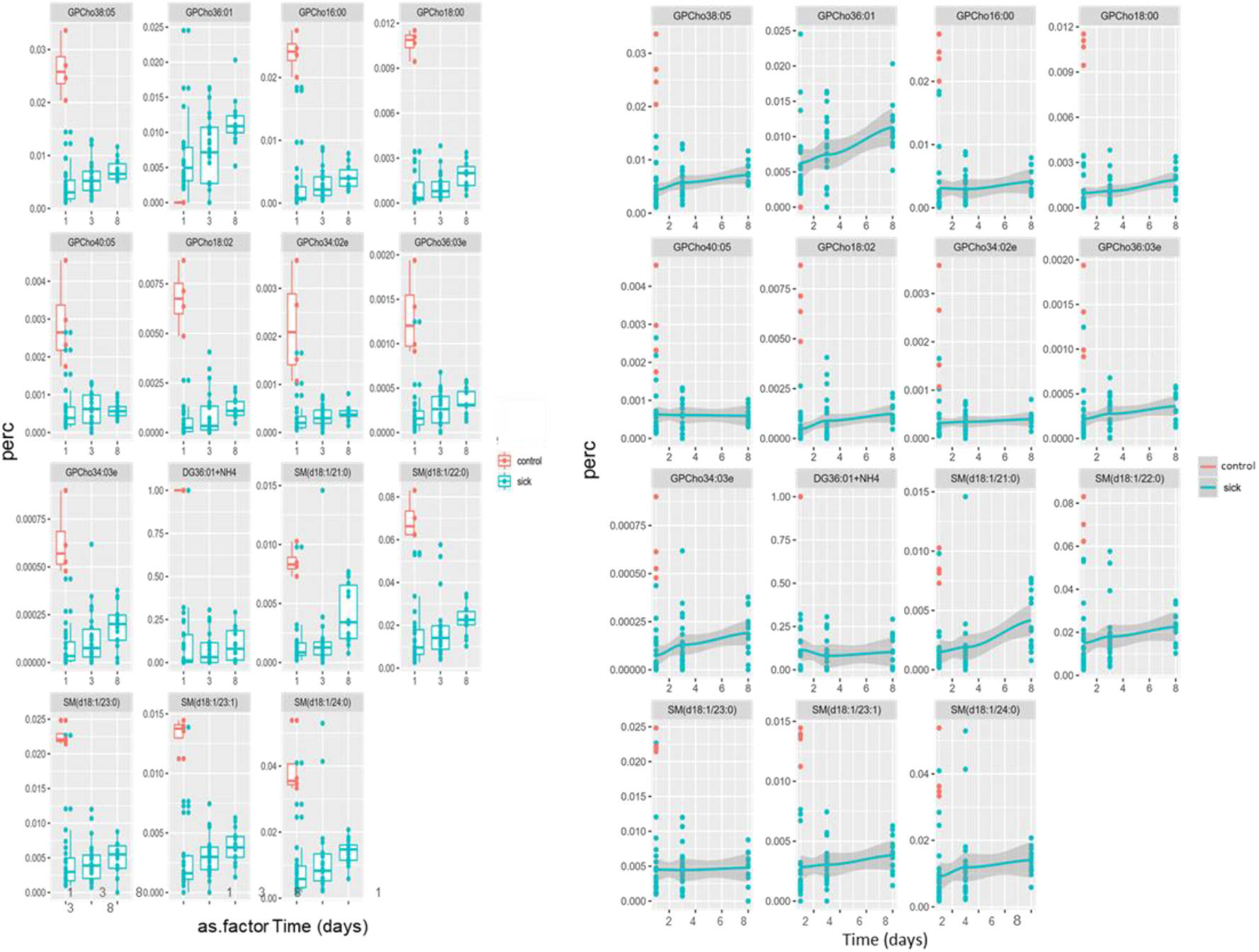
Time course for percent GPC and SM species lipids over 8 days; a-boxplots; b-geometric mean.

### 2.3 Correlations Analysis

In an attempt to better attribute the statistical differences in sphingolipid levels with physiological conditions, we ran a correlation analysis over the three time points as compared to percent total calories, percent total protein, brain injury (yes/no) and infection (bacterial/viral) (Table 3a, b; Supplemental Table 1 & 2). ECMO patients at day eight demonstrated a negative correlation to brain injury for total dhSM (−0.730), total SM (−0.730), total Hex-Cer (−0.730), and total Lac-Cer (−0.730) (Table 3a). Similarly, ECMO patients at day eight positively correlated with total protein intake for total dhSM (0.733), and total Lac-Cer (0.867), moderately with total SM (0.600), and total Hex-Cer (0.600); moderately positively correlated at day 1 for total Cer (0.600), and at day three for total 2-hydroxy Cer (0.683) (Table 3b). The sample sizes at time three for ECMO patients was small (n=6), therefore interpretation of this result must be tempered, however nutritional intake may be the most strongly correlated modifier that we explored.

**Table 3a:** Correlations between lipid values and brain injury (yes/no) in MODS patients over time

| Group | Time | Total dhSM | Total SM | Total Cer | Total hydroxy Cer | Total Hex-Cer | Total Lac-Cer |
| --- | --- | --- | --- | --- | --- | --- | --- |
| MODS | Day 1 | NA | -0.053 | -0.053 | NA | -0.172 | -0.238 |
| MODS | Day 3 | NA | -0.059 | 0.029 | -0.217 | 0.029 | 0.074 |
| MODS | Day 8 | NA | 0.071 | -0.357 | -0.218 | 0.214 | 0.214 |
| ECMO | Day 1 | -0.577 | -0.655 | -0.218 | 0.000 | -0.546 | -0.655 |
| ECMO | Day 3 | -0.149 | -0.552 | 0.138 | -0.414 | -0.276 | -0.354 |
| ECMO | Day 8 | -0.730 | -0.730 | 0.183 | -0.365 | -0.730 | -0.730 |

**Table 3b:** Kendall correlations between lipid values and percent of total protein

| Group | Time | Total dhSM | Total SM | Total Cer | Total 2-hydroxy Cer | Total Hex-Cer | Total Lac-Cer |
| --- | --- | --- | --- | --- | --- | --- | --- |
| ECMO | 1.000 | 0.499 | 0.262 | 0.577 | 0.000 | 0.157 | -0.052 |
| ECMO | 3.000 | 0.264 | 0.390 | 0.098 | 0.683 | 0.390 | -0.250 |
| ECMO | 8.000 | 0.733 | 0.600 | -0.067 | -0.067 | 0.600 | 0.867 |
| MODS | 1.000 | NA | NA | NA | NA | NA | NA |
| MODS | 3.000 | NA | NA | NA | NA | NA | NA |
| MODS | 8.000 | NA | -0.429 | -0.071 | -0.182 | -0.357 | -0.214 |

Notes: 2-hydroxy Cer: 2-hydroxy ceramide; Cer: ceramide; dhSM: dihydrosphingomyelin; Hex-Cer: hexosylceramides; Lac-Cer: lactosylceramides; SM: Sphingomyelin. Table 3a. not adjusted for sex and age.

## 3. Discussion

Sphingolipids are fundamental to the structural components of cell membranes and are involved in the regulation of biological processes including cell growth, differentiation and apoptosis. Sphingolipids were found to be lower at baseline in our patients and increased over time. In our patient population, levels of plasma Hex-Cer, a metabolite of ceramide, were found to be higher than that of sedation controls over the course of the 8 days. Lipid rafts formed through the interaction of cholesterol and glycosphingolipid, create platforms that support signal transduction, pathogen infection [23] and the viral life cycle [24]. Hex-Cer has been shown to play a role in chronic hepatitis C infections [25] and viral replication. We know that 14/24 of our study participants had infections, and we believe that our lipidomics results are linked to an infection. All of the SM’s species were low in comparison to controls which corresponds to the overall patient profiles found given by the low percentage of SP and SMs found in the critically ill cohort.

There was a large amount of variability in the glycerolipids, with time points higher than controls. Glycerolipids (mono-di-tri-glycerides) are largely exogenous in nature and obtained through dietary consumption. Most believe that TAG’s are purely for energy storage, therefore this increase in glyerolipid levels may not be surprising given the catabolic state of critically ill patients. Sepsis also causes hyper-triglyceridemia [26], which may be contributing to the spike in TAG levels. By contrast, lipotoxicity may result from an increase in TAG’s or the breakdown product of TAGs, non-esterified fatty acids (NEFA’s) [18], and TAG/TG metabolites (ceramides, diacylglycerols) [27,28].

One result was striking. GPC(36:1), which has been associated with human glioma [29], was abundant in all sick patients. One of the building blocks of GPC(36:1) is oleic acid (18:1), which is a pre-cursor to linoleic acid which feeds into the generation of arachidonic acid. Levels of oleic acid and arachidonic acid have been shown to be inversely correlated [30,31]. Some mammals lack the ability to convert linoleic acid to arachidonic acid, therefore obtaining polyunsaturated omega-6 fatty acid from dietary animal sources (meat, eggs) essential. Choline levels increase in brain tumors and inflammation. This marker may imply that our critically ill patients are undergoing some type of brain injury, beyond that which is visible by routine neurological monitoring.

In light of recent findings with COVID, the recovery from metabolic alterations can take time, as patients discharged from the hospital irrespective of severe or mild symptoms, had not yet re-normalized their metabolite and lipid levels [10]. Our data show how critical illness affect lipid metabolism, with subsequent recovery possible by dietary intervention for glycerolipids and sphingolipids.

Our study has several limitations. Large variability in the lipidomic signature of a limited number of samples is present limiting our statistical analysis; therefore we can only comment on the overall increases and decreases in the data. This was an observational study and thus we are unable to infer that changes in lipid over time are the result of clinical interventions, such as introduction of enteral feeds. Secondly, due to the many variables measured and the limited sample size, there is a possibility of Type 1 errors, and false-positive results [32], characteristic of high dimensional data anlaysis. The metabo-lome-wide significance level (MWSL) (P > 2 × 10-5 − 4 × 10-6), that should be adopted in future studies [33]. Additional confounding factors, such as pre-admission diet, medication use, medical status, and inherent metabolic and genetic differences likely exist that are not measured in this study.

## 4. Materials and Methods

### 4.1 Design, Site, Sample and Data Collection

Critically ill patients admitted to the PICU in Western Michigan, and referred by attending physician, were screened, consented, and had blood samples collected up to 3 independent time points (baseline, >48hrs, and >7days), according to IRB approval (2016-062-SH/HDVCH), as previously described [18,19,34,35]. If patients were discharged before later time points, samples were not collected. Helen DeVos Children’s Hospital PICU sees 1500 admissions per year (over 6000 patient days), has seventeen board certified intensivists covering a 24-bed unit with flex capability to care for up to 36 critically ill children, including a cardiothoracic surgery program and a level 1 trauma center. Patients with MODS fit the following inclusion criteria: < 18 years of age; on vasopressors with a central line and requiring respiratory support. Sedation control patients were presenting for routine sedation with benign characteristics. Patients were excluded if: they had a known auto-immune disease, had a limitation of care in their advance directives, had undergone cardiopulmonary bypass prior to onset of MODS, received plasmapheresis prior to ECMO initiation, and were patients of the neonatal intensive care unit. Blood samples were drawn in EDTA-filled anticoagulant, centrifuged and stored at −80°C. Basic demographic variables were pulled from the local electronic medical record (EMR). Dietary history was extracted from the dietician’s EMR, as previously reported [18]. Data was collected and managed using REDCap [36]. Severity of illness scores were retrieved through the Virtual Pediatric Intensive Care Unit Performance Systems (VPS, LLC, Los Angeles, CA). Nutritional intake was previously described [18].

### 4.2 Methods

Lipidome profiles were determined from plasma subjected to lipid extraction with, acetone, methanol, and acetonitrile [37], as previously described [18]. Extracts were analyzed by nano-electrospray (nESI) direct infusion high resolution/accurate mass spectrometry (MS) and tandem mass spectrometry (MS/MS) utilizing an LTQ-Orbitrap Velos mass spectrometer (Thermo Scientific, Waltham, MA). An Advion Nanomate Triversa (Advion Biosciences, Ithaca, NY) served as the nESI source and high throughput autosampler. Data were acquired for two minutes for each MS analysis. For MS/MS verification of lipid identities, Higher-Energy Collisional Dissociation (HCD) was utilized with the FT analyzer operating at 100,000 resolving power in order to obtain accurate mass measurements of lipid product ions and increase confidence in product ion assignments. Lipidome analysis provided untargeted analysis across all classes of glycerolipids (GC) (including mono-, di- and triglycerides: DAG’s, TAG’s), glycerphospholipids (GP), sphingolipids (SP), for this report (other lipid classes were previously reported [18]). Additional species analysis was completed on phospholipids, triacylglycerides (TAG’s), di-acylglycerides (DAG’s), cholesterol (chol), and sphingomyelins (SM). Di-myristoyl phosphatidylcholine served as an internal standard. Limits of detection ranged from 0.01 nM to 10 nM across all lipid classes. Each sample was run twice (positive and negative ion analysis) and data were combined. Peak findings and quantification was performed with Lipid Mass Spectrum Analysis (LIMSA) version 1.0 software [38].

### 4.3 Analysis

The decision was made a priori to compare the sedation control population (n = 4) with all of the critically ill children (n = 24), unless stated otherwise. Some statistical analyses were performed using R (v 3.6.0) [39]. Where appropriate, percent data were transformed before being analyzed with a beta regression from the R package betareg [40], and MedCalc (MedCalc Software Ltd, Ostend, Belgium). Total normalized ion values were log-transformed and then analyzed using generalized linear regression models (glm) [41]. All regression models were adjusted for age and sex. Contrasts between treatment groups (sick vs. sedation) were conducted using R package emmeans [42]. P-values from regression analyses have been corrected for multiple testing via the FDR method. We focused on glycerol-glycerophospho- and sphingo-lipids. Additionally, independent t-tests, Welch’s t-tests, and F-tests of equal variances were run on these data and included multiple testing adjustment. Correlation analysis was performed where indicated using Kendall Tau, whereby percent calories and protein intake were calculated from resting energy expenditure [43], and daily required intake [44] (≤ 33 %; 34 % - 66; and ≥ 67 %), binary values (yes/no) for brain injury and infection (bacterial/viral).

## 5. Conclusions

We note low abundance of sphingolipids and sphingomyelins for this cohort of critically-ill pediatric patients. Trace amounts of markers that are pro-apoptotic and associated with membrane homeostasis were also found. A ceramide metabolite previously associated with infection Hex-Cer, was elevated, in concert with Lac-Cer an intermediary in glycosphingolipid metabolism may have a protective effect against infection. In total, our observations display a complex interplay of lipids, with a lipid species of GPC(36:1) highlighted as a potential biomarker candidate of critical illness. The pathways for biosynthesis of these lipids are well known, future work will involve exploration of these pathways.

## Data Availability

Data is available upon request.

## 6. Abbreviations

Cer: ceramide
chol: cholesterol
DAG’s: diacylglycerides
dhSM: dihydro sphingomyelin
ECMO: extracorporeal membrane oxygenation
EMR: electronic medi cal record
FA’s: fatty acids
GC: glycerolipids
GP: glycerophospholipids
GPC: glycerophosphatidylcholine
Hex-Cer: hexosylceramides
HLH: hemoph agocytic lymphohistiocytosis
2-hydroxy Cer: 2-hydroxy ceramide
Lac-Cer: lacto sylceramides
MODS: multi-organ dysfunction syndrome
NADPH: nicotinamide adenine dinucleotide phosphate
NEFA’s: non-esterified fatty acids
NPO: nil per os
PICU: Pediatric Intensive Care Unit
PICU LOS: PICU length of stay
PELOD: Pediatric Logistic Organ Dysfunction-2 score
SD: standard deviation
SM: sphingomye lins
SP: sphin golipids
TAG’s: triacylglycerides.

## Supplementary Materials

**Table S1**. Kendall Correlations between Lipid Value and Percent of Total Calories; **Table S2:** Correlations between lipid values and viral infection (yes/no) in patients over time; not adjusted for age or sex; **Figure S1**. Percent total cholesterol lipid species; **Figure S2**. Percent total triacylglycerol lipid species; **Figure S3**. Percent total diacylglycerol lipid species.

## Funding

Funding for this project was provided for SR, ML by Spectrum Health Office of Research (SHOR) funding initiative for precision medicine (#R51100431217), Helen DeVos Children’s Hospital Foundation (HDVCH), grant (#R51100881018), and HDVCH Foundation grant (DG, SR, ML). NIH Office of the Director and NIEHS grant K01ES025435 (JWP).

## Institutional Review Board Statement

The study was conducted according to the guidelines of the Declaration of Helsinki, and approved by the Institutional Review Board (or Ethics Committee) of Spectrum Health (protocol code IRB 2016-062-SH/HDVCH).

## Informed Consent Statement

Informed consent was obtained from all subjects involved in the study.

## Acknowledgments

The authors would like to thank the PICU staff at Helen DeVos Children’s Hospital for their support in the completion of this study and various contributions, and Brittany Essenmacher for the critical review of the final manuscript.

## Conflicts of Interest

The authors declare no conflict of interest. The funders had no role in the design of the study; in the collection, analyses, or interpretation of data; in the writing of the manuscript, or in the decision to publish the results.

## Notes

### Competing Interest Statement

The authors have declared no competing interest.

### Author Declarations

The study was conducted according to the guidelines of the Declaration of Helsinki, and ap-proved by the Institutional Review Board (or Ethics Committee) of Spectrum Health (protocol code IRB 2016-062-SH/HDVCH).

## References

1. Quehenberger, O., Dennis, E.A. The human plasma lipidome. The New England journal of medicine 2011, 365, 1812–1823, doi:10.1056/NEJMra1104901.

2. Dennis, E.A. Lipidomics joins the omics evolution. Proceedings of the National Academy of Sciences of the United States of America 2009, 106, 2089–2090, doi:10.1073/pnas.0812636106.

3. Typpo, K.V., Petersen, N.J., Hallman, D.M., Markovitz, B.P., Mariscalco, M.M. Day 1 multiple organ dysfunction syndrome is associated with poor functional outcome and mortality in the pediatric intensive care unit. Pediatr Crit Care Med 2009, 10, 562–570, doi:10.1097/PCC.0b013e3181a64be1.

4. Typpo, K., Watson, R.S., Bennett, T.D., Farris, R.W.D., Spaeder, M.C., Petersen, N.J., Pediatric Existing Data Analysis, I., Pediatric Acute Lung, I., Sepsis Investigators, N. Outcomes of Day 1 Multiple Organ Dysfunction Syndrome in the PICU. Pediatr Crit Care Med 2019, 20, 914–922, doi:10.1097/PCC.0000000000002044.

5. Mickiewicz, B., Vogel, H.J., Wong, H.R., Winston, B.W. Metabolomics as a novel approach for early diagnosis of pediatric septic shock and its mortality. Am J Respir Crit Care Med 2013, 187, 967–976, doi:10.1164/rccm.201209-1726OC.

6. Wong, H.R., Cvijanovich, N., Lin, R., Allen, G.L., Thomas, N.J., Willson, D.F., Freishtat, R.J., Anas, N., Meyer, K., Checchia, P.A., et al. Identification of pediatric septic shock subclasses based on genome-wide expression profiling. BMC Med 2009, 7, 34, doi:10.1186/1741-7015-7-34.

7. Wong, H.R., Cvijanovich, N.Z., Allen, G.L., Thomas, N.J., Freishtat, R.J., Anas, N., Meyer, K., Checchia, P.A., Lin, R., Shanley, T.P., et al. Validation of a gene expression-based subclassification strategy for pediatric septic shock. Crit Care Med 2011, 39, 2511–2517, doi:10.1097/CCM.0b013e3182257675.

8. Langley, R.J., Tsalik, E.L., van Velkinburgh, J.C., Glickman, S.W., Rice, B.J., Wang, C., Chen, B., Carin, L., Suarez, A., Mohney, R.P., et al. An integrated clinico-metabolomic model improves prediction of death in sepsis. Science translational medicine 2013, 5, 195ra195, doi:10.1126/scitranslmed.3005893.

9. Barbaro, R.P., Paden, M.L., Guner, Y.S., Raman, L., Ryerson, L.M., Alexander, P., Nasr, V.G., Bembea, M.M., Rycus, P.T., Thiagarajan, R.R. Pediatric Extracorporeal Life Support Organization Registry International Report 2016. ASAIO journal (American Society for Artificial Internal Organs : 1992) 2017, 63, 456–463, doi:10.1097/mat.0000000000000603.

10. Wu, D., Shu, T., Yang, X., Song, J.-X., Zhang, M., Yao, C., Liu, W., Huang, M., Yu, Y., Yang, Q., et al. Plasma Metabolomic and Lipidomic Alterations Associated with COVID-19. National Science Review 2020, 10.1093/nsr/nwaa086, doi:10.1093/nsr/nwaa086.

11. Kyle, J.E., Burnum-Johnson, K.E., Wendler, J.P., Eisfeld, A.J., Halfmann, P.J., Watanabe, T., Sahr, F., Smith, R.D., Kawaoka, Y., Waters, K.M., et al. Plasma lipidome reveals critical illness and recovery from human Ebola virus disease. Proc Natl Acad Sci U S A 2019, 116, 3919–3928, doi:10.1073/pnas.1815356116.

12. Briassoulis, G., Venkataraman, S., Thompson, A. Cytokines and metabolic patterns in pediatric patients with critical illness. Clin Dev Immunol 2010, 2010, 354047, doi:10.1155/2010/354047.

13. Maile, M.D., Standiford, T.J., Engoren, M.C., Stringer, K.A., Jewell, E.S., Rajendiran, T.M., Soni, T., Burant, C.F. Associations of the plasma lipidome with mortality in the acute respiratory distress syndrome: a longitudinal cohort study. Respir Res 2018, 19, 60–60, doi:10.1186/s12931-018-0758-3.

14. Erion, D.M., Shulman, G.I. Diacylglycerol-mediated insulin resistance. Nature medicine 2010, 16, 400–402, doi:10.1038/nm0410-400.

15. Jenkins, R.W., Clarke, C.J., Lucas, J.T., Jr., Shabbir, M., Wu, B.X., Simbari, F., Mueller, J., Hannun, Y.A., Lazarchick, J., Shirai, K. Evaluation of the role of secretory sphingomyelinase and bioactive sphingolipids as biomarkers in hemophagocytic lymphohistiocytosis. Am J Hematol 2013, 88, E265–272, doi:10.1002/ajh.23535.

16. Koal, T., Klavins, K., Seppi, D., Kemmler, G., Humpel, C. Sphingomyelin SM(d18:1/18:0) is significantly enhanced in cerebrospinal fluid samples dichotomized by pathological amyloid-beta42, tau, and phospho-tau-181 levels. J Alzheimers Dis 2015, 44, 1193–1201, doi:10.3233/JAD-142319.

17. Nixon, G.F. Sphingolipids in inflammation: pathological implications and potential therapeutic targets. British journal of pharmacology 2009, 158, 982–993, doi:10.1111/j.1476-5381.2009.00281.x.

18. Leimanis-Laurens, M.L., Ferguson, K., Wolfrum, E., Boville, B., Sanfilippo, D., Lydic, T., Prokop, J., Rajasekaran, S. Pediatric Multi-Organ Dysfunction Syndrome: Analysis by an Untargeted Shotgun Lipidomic Approach Reveals Low-abundance Plasma Phospholipids and Dynamic Recovery Over 8-Day Period, a Single-Center Observational Study. medRxiv 2020, 10.1101/2020.11.24.20237891, 2020.2011.2024.20237891, doi:10.1101/2020.11.24.20237891.

19. Shankar, R., Leimanis, M.L., Newbury, P.A., Liu, K., Xing, J., Nedveck, D., Kort, E.J., Prokop, J.W., Zhou, G., Bachmann, A.S., et al. Gene expression signatures identify paediatric patients with multiple organ dysfunction who require advanced life support in the intensive care unit. EBioMedicine 2020, 62, 103122, doi:https://doi.org/10.1016/j.ebiom.2020.103122.

20. Vieira, C.R., Munoz-Olaya, J.M., Sot, J., Jiménez-Baranda, S., Izquierdo-Useros, N., Abad, J.L., Apellániz, B., Delgado, R., Martinez-Picado, J., Alonso, A., et al. Dihydrosphingomyelin Impairs HIV-1 Infection by Rigidifying Liquid-Ordered Membrane Domains. Chemistry & Biology 2010, 17, 766–775, doi:https://doi.org/10.1016/j.chembiol.2010.05.023.

21. Kota, V., Hama, H. 2’-Hydroxy ceramide in membrane homeostasis and cell signaling. Adv Biol Regul 2014, 54, 223–230, doi:10.1016/j.jbior.2013.09.012.

22. Muilwijk, M., Goorden, S.M.I., Celis-Morales, C., Hof, M.H., Ghauharali-van der Vlugt, K., Beers-Stet, F.S., Gill, J.M.R., Vaz, F.M., van Valkengoed, I.G.M. Contributions of amino acid, acylcarnitine and sphingolipid profiles to type 2 diabetes risk among South-Asian Surinamese and Dutch adults. BMJ open diabetes research & care 2020, 8, doi:10.1136/bmjdrc-2019-001003.

23. van der Meer-Janssen, Y.P.M., van Galen, J., Batenburg, J.J., Helms, J.B. Lipids in host-pathogen interactions: pathogens exploit the complexity of the host cell lipidome. Progress in lipid research 2010, 49, 1–26, doi:10.1016/j.plipres.2009.07.003.

24. Negro, F. Abnormalities of lipid metabolism in hepatitis C virus infection. Gut 2010, 59, 1279–1287, doi:10.1136/gut.2009.192732.

25. Zhang, J.-Y., Qu, F., Li, J.-F., Liu, M., Ren, F., Zhang, J.-Y., Bian, D.-D., Chen, Y., Duan, Z.-P., Zhang, J.-L., et al. Up-regulation of Plasma Hexosylceramide (d18: 1/18: 1) Contributes to Genotype 2 Virus Replication in Chronic Hepatitis C: A 20-Year Cohort Study. Medicine (Baltimore) 2016, 95, e3773–e3773, doi:10.1097/MD.0000000000003773.

26. Caspar-Bauguil S., Genestal M. Plasma Phospholipid Fatty Acid Profiles in Septic Shock. In Diet and Nutrition in Critical Care, R., R., V.R., P., V.B., P., Eds. Springer: New York, NY, 2015; DOI 10.1007/978-1-4614-7836-2_137p. 219.

27. Schaffer, J.E. Lipotoxicity: when tissues overeat. Curr Opin Lipidol 2003, 14, 281–287, doi:10.1097/00041433-200306000-00008.

28. Listenberger, L.L., Han, X., Lewis, S.E., Cases, S., Farese, R.V., Jr., Ory, D.S., Schaffer, J.E. Triglyceride accumulation protects against fatty acid-induced lipotoxicity. Proc Natl Acad Sci U S A 2003, 100, 3077–3082, doi:10.1073/pnas.0630588100.

29. Li, W., Jia, H., Li, Q., Cui, J., Li, R., Zou, Z., Hong, X. Glycerophosphatidylcholine PC(36:1) absence and 3’-phosphoadenylate (pAp) accumulation are hallmarks of the human glioma metabolome. Scientific reports 2018, 8, 14783, doi:10.1038/s41598-018-32847-8.

30. Høstmark, A.T., Haug, A. Percentages of oleic acid and arachidonic acid are inversely related in phospholipids of human sera. Lipids in health and disease 2013, 12, 106–106, doi:10.1186/1476-511X-12-106.

31. Høstmark, A.T., Haug, A. The inverse association between relative abundances of oleic acid and arachidonic acid is related to alpha-linolenic acid. Lipids in Health and Disease 2014, 13, 76, doi:10.1186/1476-511X-13-76.

32. Han, X., Rozen, S., Boyle, S.H., Hellegers, C., Cheng, H., Burke, J.R., Welsh-Bohmer, K.A., Doraiswamy, P.M., Kaddurah-Daouk, R. Metabolomics in early Alzheimer’s disease: identification of altered plasma sphingolipidome using shotgun lipidomics. PLoS One 2011, 6, e21643, doi:10.1371/journal.pone.0021643.

33. Chadeau-Hyam, M., Ebbels, T.M., Brown, I.J., Chan, Q., Stamler, J., Huang, C.C., Daviglus, M.L., Ueshima, H., Zhao, L., Holmes, E., et al. Metabolic profiling and the metabolome-wide association study: significance level for biomarker identification. J Proteome Res 2010, 9, 4620–4627, doi:10.1021/pr1003449.

34. Prokop, J.W., Shankar, R., Gupta, R., Leimanis, M.L., Nedveck, D., Uhl, K., Chen, B., Hartog, N.L., Van Veen, J., Sisco, J.S., et al. Virus-induced genetics revealed by multidimensional precision medicine transcriptional workflow applicable to COVID-19. Physiological genomics 2020, 52, 255–268, doi:10.1152/physiolgenomics.00045.2020.

35. Leimanis-Laurens, M.L., Gil, D., Kampfshulte, A., Krohn, C., Prentice, E., Sanfilippo, D., Prokop, J.W., Lydic, T., Rajasekaran, S. The Feasibility of Studying Metabolites in PICU Multi-Organ Dysfunction Syndrome Patients Over an 8-day Course Using An Untargeted Approach. medRxiv 2020, 10.1101/2020.12.04.20244053, 2020.2012.2004.20244053, doi:10.1101/2020.12.04.20244053.

36. Harris, P.A., Taylor, R., Thiekle, R., Payne, J., Gonzalez, N., Conde, J.G. Research electronic capture (REDCap) - A metadata-driven methodology and workflow process for providing translational research informatics supports. Journal of Biomedical Information 2009, 42, 377–381.

37. Cai, X., Li, R. Concurrent profiling of polar metabolites and lipids in human plasma using HILIC-FTMS. Scientific reports 2016, 6, 36490, doi:10.1038/srep36490.

38. Haimi, P., Uphoff, A., Hermansson, M., Somerharju, P. Software tools for analysis of mass spectrometric lipidome data. Anal Chem 2006, 78, 8324–8331, doi:10.1021/ac061390w.

39. Team, R.C. R: A language and environment for statistical computing., Vienna, Austria., 2019.

40. Cribari-Neto, F., Zeileis, A. Beta Regression in R. Journal of Statistical Software 2010, 34, 1–24.

41. Bates, D., Maehler, M., Bolker, B., Walker, S. Fitting Linear Mixed-Effects Models Using lme4. Journal of Statistical Software 2015, 67, 1–48, doi:10.18637/jss.v067.i01.

42. Lenth, R. Emmeans: Estimated Marginal Means, aka Least-Squares Means. R package version 1.4 2019.

43. Schofield, W.N. Predicting basal metabolic rate, new standards and review of previous work. Human nutrition. Clinical nutrition 1985, 39 Suppl 1, 5–41.

44. Medicine, I.o. Dietary Reference Intakes: The Essential Guide to Nutrient Requirements; The National Academies Press: Washington, DC, 2006; doi:10.17226/11537 pp. 1344.

