## Supplemental for "Hexosylceramides and glycerophosphatidylcholine GPC(36:1) increase in Multi-Organ Dysfunction Syndrome patients with Pediatric Intensive Care Unit Admission over 8-day hospitalization"

Article

| **Supplemental Table 1.** Kendall Correlations between Lipid Value and Percent of Total Calories | | | | | | | |
| --- | --- | --- | --- | --- | --- | --- | --- |
| Group | Time | Total dhSM | Total SM | Total Cer | Total 2-hydroxy  Cer | Total Hex-Cer | Total Lac-Cer |
| ECMO | Day 1 | 0.499 | 0.262 | 0.577 | 0.000 | 0.157 | -0.052 |
| ECMO | Day 3 | 0.158 | 0.293 | 0.000 | 0.586 | 0.293 | -0.350 |
| ECMO | Day 8 | 0.200 | -0.200 | 0.467 | -0.333 | 0.067 | 0.067 |
| MODS | Day 1 | N/A | N/A | N/A | N/A | N/A | N/A |
| MODS | Day 3 | N/A | N/A | N/A | N/A | N/A | N/A |
| MODS | Day 8 | N/A | -0.429 | -0.214 | -0.182 | -0.214 | -0.071 |
| Notes: 2-hyroxy Cer: 2-hydroxy ceramide; Cer: ceramide; dhSM: dihydrosphingomyelin; Hex-Cer: hexosylceramides; Lac-Cer: lactosylceramides; SM: Sphingomyelin. | | | | | | | |

| **Supplemental Table 2:** Correlations between lipid values and viral infection (yes/no) in patients over time; not adjusted for age or sex | | | | | | | |
| --- | --- | --- | --- | --- | --- | --- | --- |
| Group | Time | Infection | Total dhSM | Total SM | Total Cer | Total hydroxy-Cer | Total Lac-Cer |
| MODS | Day 1 | Bacterial | N/A | 0.024 | 0.189 | N/A | 0.450 |
| MODS | Day 3 | Bacterial | N/A | -0.159 | -0.106 | 0.447 | 0.080 |
| MODS | Day 8 | Bacterial | N/A | -0.436 | -0.109 | 0.111 | -0.109 |
| MODS | Day 1 | Viral | N/A | -0.382 | -0.209 | N/A | -0.247 |
| MODS | Day 3 | Viral | N/A | 0.559 | 0.206 | -0.309 | 0.118 |
| MODS | Day 8 | Viral | N/A | 0.000 | -0.109 | 0.056 | 0.000 |
| ECMO | Day 1 | Viral | 0.361 | 0.049 | -0.049 | -0.447 | -0.146 |
| ECMO | Day 3 | Viral | 0.136 | 0.252 | 0.504 | 0.378 | -0.065 |
| ECMO | Day 8 | Viral | -0.086 | -0.086 | -0.086 | 0.430 | 0.086 |

Notes: 2-hyroxy Cer: 2-hydroxy ceramide; Cer: ceramide; dhSM: dihydrosphingomyelin; Hex-Cer: hexosylceramides; Lac-Cer: lactosylceramides; SM: Sphingomyelin.

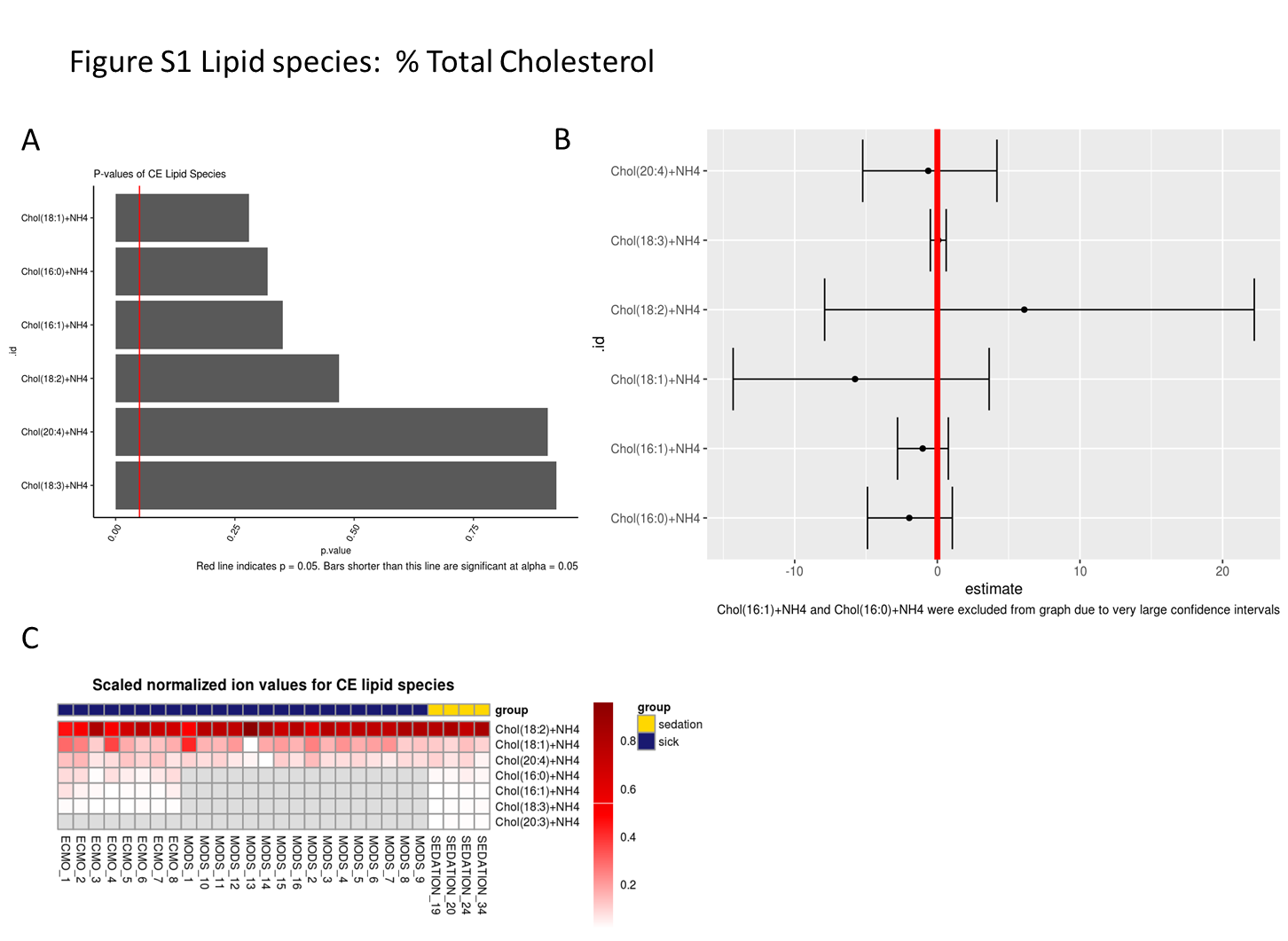
**Figure S1.** Percent total cholesterol lipid species

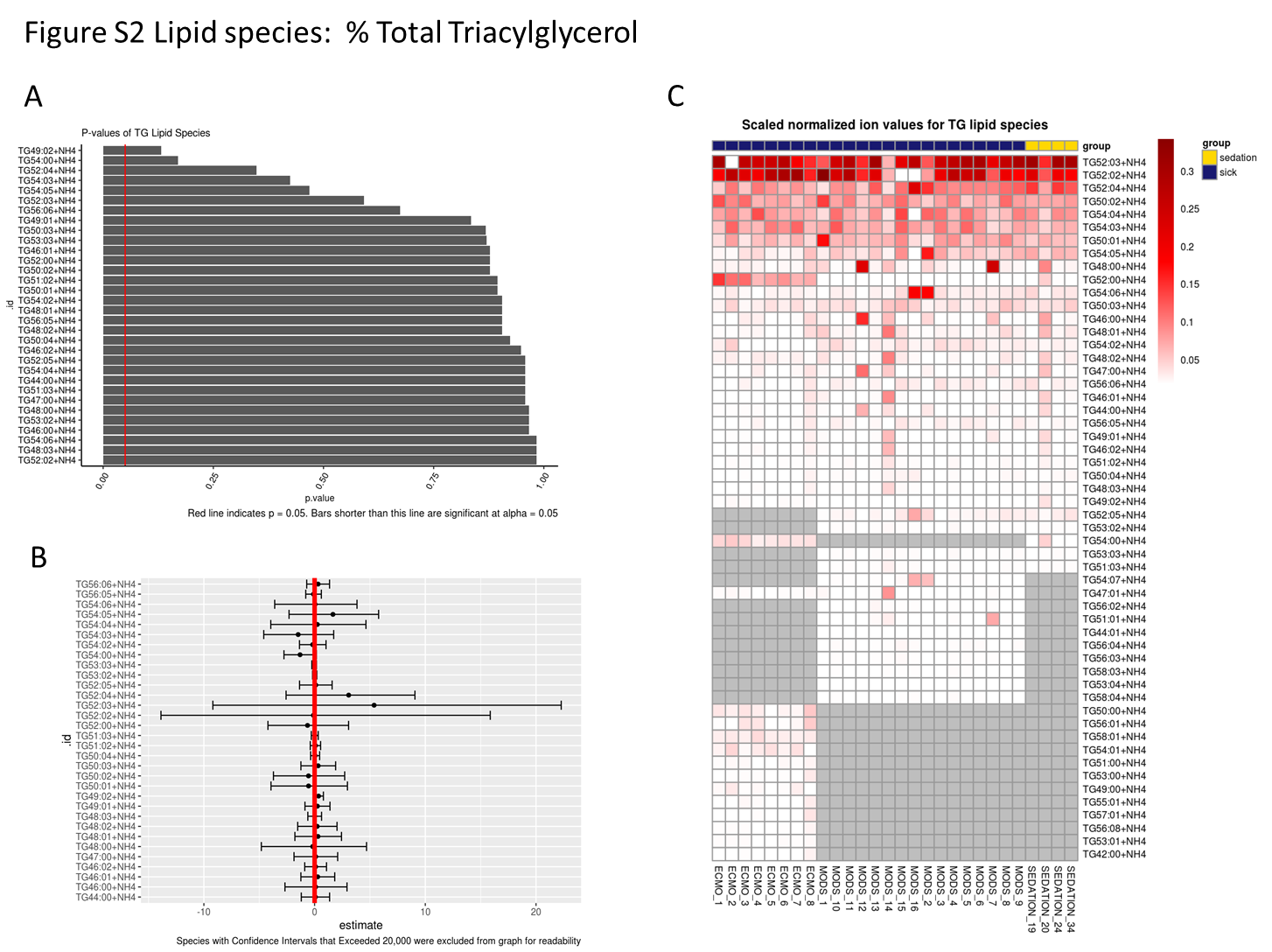
**Figure S2.** Percent total triacylglycerol lipid species

**Figure S3.** Percent total diacylglycerol lipid species

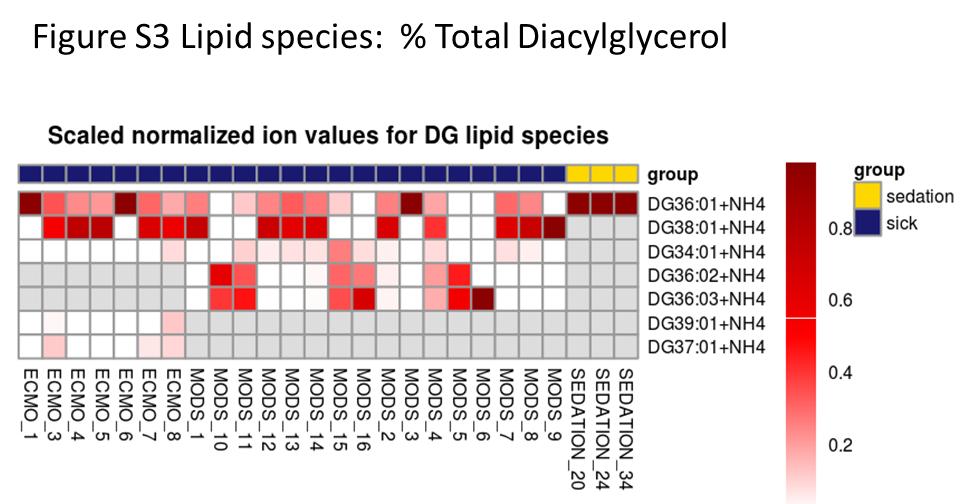
